# Plasma cell-free DNA promise disease monitoring and tissue injury assessment of COVID-19

**DOI:** 10.1101/2021.07.19.21260139

**Authors:** Xin Jin, Yanqun Wang, Jinjin Xu, Yimin Li, Fanjun Cheng, Yuxue Luo, Haibo Zhou, Shanwen Lin, Fei Xiao, Lu Zhang, Yu Lin, Zhaoyong Zhang, Yan Jin, Fang Zheng, Wei Chen, Airu Zhu, Ye Tao, Jingxian Zhao, Tingyou Kuo, Yuming Li, Lingguo Li, Liyan Wen, Rijing Ou, Fang Li, Long Lin, Yanjun Zhang, Jing Sun, Hao Yuan, Zhen Zhuang, Haixi Sun, Zhao Chen, Jie Li, Jianfen Zhuo, Dongsheng Chen, Shengnan Zhang, Yuzhe Sun, Peilan Wei, Jinwei Yuan, Tian Xu, Huanming Yang, Jian Wang, Xun Xu, Nanshan Zhong, Yonghao Xu, Kun Sun, Jincun Zhao

## Abstract

COVID-19 is a huge threat to global health. Due to the lack of definitive etiological therapeutics currently, effective disease monitoring is of high clinical value for better healthcare and management of the large number of COVID-19 patients. In this study, we recruited 37 COVID-19 patients, collected 176 blood samples upon diagnosis and during treatment, and analyzed cell-free DNA (cfDNA) in these samples. We report gross abnormalities in cfDNA of COVID-19 patients, including elevated GC content, altered molecule size and end motif patterns. More importantly, such cfDNA characteristics reflect patient-specific physiological conditions during treatment. Further analysis on tissue origin tracing of cfDNA reveals frequent tissue injuries in COVID-19 patients, which is supported by clinical diagnoses. Hence, we demonstrate the translational merit of cfDNA as valuable analyte for effective disease monitoring, as well as tissue injury assessment in COVID-19 patients.

## Introduction

The current pandemic, COVID-19, has become a huge threat to global health: as many as 130 million patients had been diagnosed worldwide by early-Apr 2021. At present, there is no effective etiological treatment for COVID-19, and the number of diagnosed patients increases rapidly. Considering the tremendous amount of COVID-19 patients, disease monitoring is of particular clinical value for better management of these patients (1); however, efficient approaches are still limited. Since the pathogen of COVID-19 is a coronavirus named SARS-CoV-2 (2), nucleic acid test of the pathogenic virus has become a standard method for diagnosis, treatment monitoring and cure (3–5). Most patients turn negative for the nucleic acid test of SARS-CoV-2 several weeks after treatment, but some patients show persistent viral shedding with low IgG antibody response. Considering that many asymptomatic and discharged patients are also positive for SARS-CoV-2 test, additional diagnostic approaches are thus needed for better disease monitoring of the patients. Furthermore, as evidenced in multiple investigations (6–11), COVID-19 causes damages to various organs including lungs, the primary infected tissue, as well as heart, kidney, and brain. Such damage could further induce organ failures, shock, acute respiratory distress syndrome and even patient mortality. Hence, to take early intervention measurements to prevent the occurrence of serious complications of the patients, it is of particular importance and urgent clinical need to develop methods for disease monitoring and organ injury assessment of COVID-19 patients.

Plasma circulating cell-free DNA (cfDNA) in peripheral blood has been discovered and actively studied for more than 70 years (12). CfDNA molecules are mostly derived from dying cells and retain various cell-type-specific signatures (13–16). Numerous studies have demonstrated that cfDNA is a valuable analyte for diagnosis and monitoring of various diseases (17, 18). In healthy subjects, cfDNA mostly originate from the hematopoietic system (19, 20); while in organ transplantation and cancer patients, cfDNA molecules released from the affected organs are readily detectable (21, 22). Moreover, cfDNA molecules are rapidly cleared with a short half-life time (typically a few hours) (23), thus reflect the real-time responses of the human body. The successful applications of plasma cfDNA in various physiological and pathological conditions suggest that these molecules may promise a practical and efficient approach for COVID-19 monitoring; however, comprehensive investigations have not been explored yet.

In this study, we have utilized plasma cfDNA to investigate the disease dynamics of COVID-19 patients during treatment. We have collected and analyzed a total of 208 blood samples from 37 COVID-19 patients and 32 controls. We report gross abnormalities, dynamics as well as organ injury signals in cfDNA, demonstrating high clinical potential of these analytes for effective disease monitoring and tissue injury assessment of COVID-19.

## Results

### Overview of the study

Fig. 1 shows the overall design of this study. A total of 37 COVID-19 patients, either in mild (N=18) or severe (N=19) conditions, were recruited from local hospitals in Guangdong province of China. Table 1 summarizes the major clinical characteristics of these patients. Briefly, in the COVID-19 patients, severe cases suffer from acute severe viral pneumonia and show serious clinical symptoms that require mechanical ventilation and intensive care unit treatment, while mild cases show weak symptoms of pneumonia (usually minor upper respiratory tract infection) and recover within a few weeks (24–27). All the COVID-19 patients are immediately hospitalized upon diagnosis; for all COVID-19 patients, the first blood-collection timepoints are within 3 days after diagnosis. All COVID-19 patients receive standard treatment following the “Diagnosis and Treatment Protocol for Novel Coronavirus Pneumonia (Trial Version 5)” guidelines published by National Health Commission & National Administration of Traditional Chinese Medicine of China. In short, all COVID-19 patients receive antiviral treatment; severe patients receive additional antibacterial treatment, and most of them also receive antifungal treatment (Table 1). Notably, 1 severe patient also receives convalescent plasma therapy (Focosi et al. 2020; National Health Commission & National Administration of Traditional Chinese Medicine 2020) on day 16 of hospitalization. The most common comorbidity in the COVID-19 patients is hypertension (4 and 6 in mild and severe groups, respectively), followed by type-II diabetes (Table 1). A total of 176 blood samples were collected at multiple timepoints upon hospitalization and during treatment. In addition, 32 age-matched non-COVID-19 controls were also recruited. CfDNA from all blood samples were investigated. Key clinical data, including SARS-CoV-2-specific immunoglobulin (i.e., IgG and IgM) levels, Chest X-ray, Computed Tomography (CT) scan, coagulation profile, liver and renal functions, electrolyte, myocardial enzymes, interleukin-6, TNF-α, procalcitonin and C-reactive protein levels, were also collected (when available) during treatment to analyze the disease states of the patients. The plasma cfDNA was extracted, sequenced, and analyzed to investigate their correlations with COVID-19 as well as dynamics during treatment.

**Figure 1.**
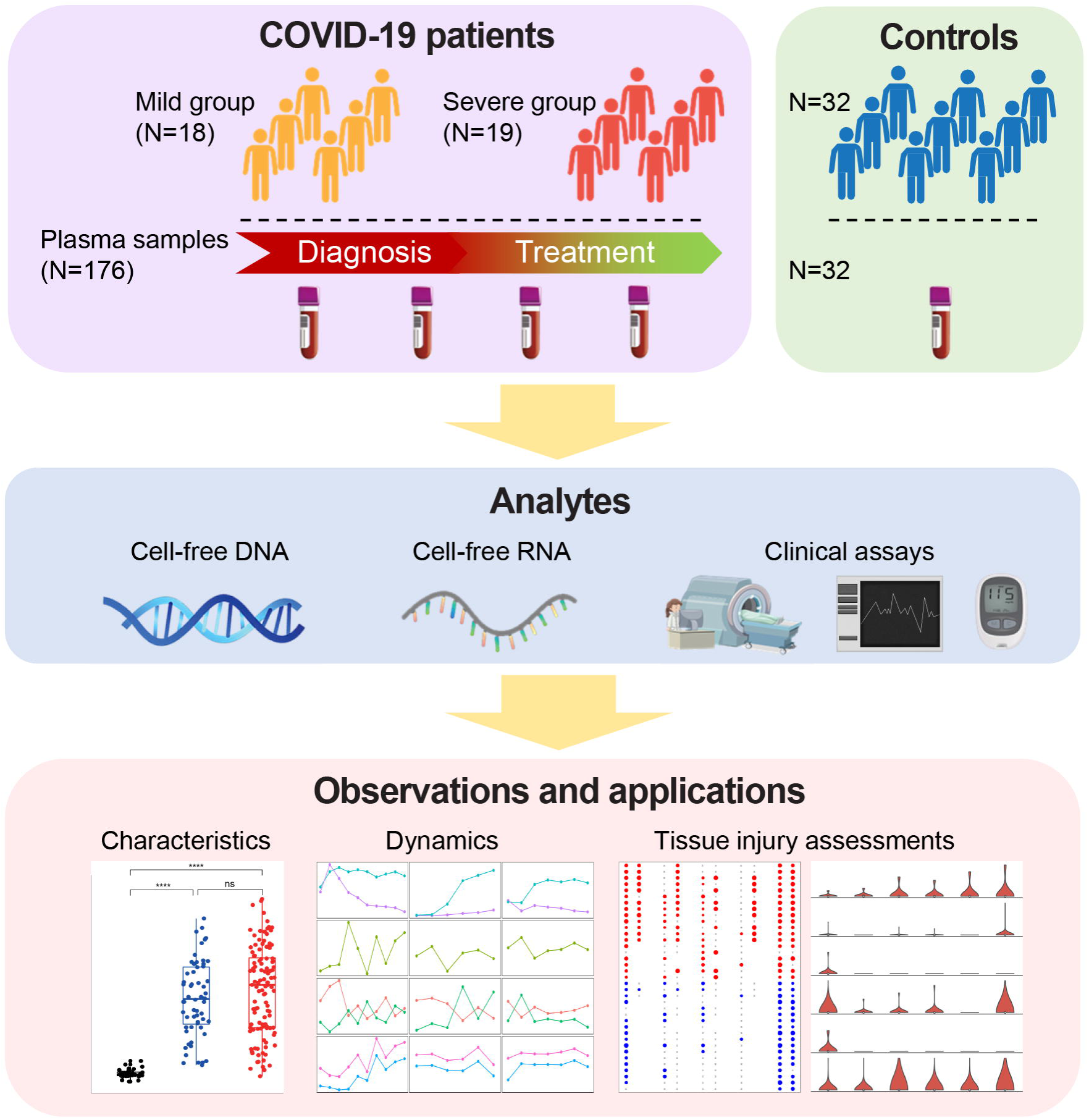
Overview of the study. A total of 37 COVID-19 patients (18 and 19 in mild severe conditions, respectively) and 32 healthy controls were recruited in this study. For the COVID-19 patients, 176 blood samples are collected upon hospitalization and during treatment. Plasma cfDNA is extracted and analyzed together with clinical data. As a result, we report disease-specific characteristics, dynamics, and tissue injury signals in cfDNA of COVID-19 patients.

**Table 1.** Characteristics of COVID-19 patients and controls.

| **Demographics** | **Controls** | **Mild patients** | **Severe patients** |
| --- | --- | --- | --- |
| Number | 32 | 18 | 19 |
| Age (years)* | 45.61 ($\pm 8.07$) | 49.61 ($\pm 18.53$) | 59.16 ($\pm 13.82$) |
| Female (proportion) | 3 (9.38%) | 7 (38.89%) | 4 (21.05%) |
| **Comorbidities** |  |  |  |
| Hypertension | 0 | 4 | 6 |
| Type 2 diabetes mellitus | 0 | 1 | 5 |
| Cardiovascular disease | 0 | 0 | 3 |
| Gallbladder disease | 0 | 0 | 3 |
| Chronic obstructive pulmonary disease | 0 | 0 | 2 |
| Hepatitis B virus infection | 0 | 0 | 2 |
| Metabolic arthritis | 0 | 1 | 1 |
| Kidney cysts | 0 | 0 | 2 |
| Thalassemia | 0 | 1 | 0 |
| **Medications** |  |  |  |
| Antiviral | NA | 18 | 19 |
| Antibacterial | NA | 3 | 19 |
| Antifungal | NA | 0 | 16 |
| Glucocorticoid | NA | 0 | 6 |

\* Age is presented as mean ( ± sd).

### Abnormalities in cfDNA of COVID-19 patients

In a previous work, we found that SARS-CoV-2-derived DNA does not present in the plasma of COVID-19 patients (which is in consistent with the RNA-virus nature of SARS-CoV-2) (28); hence, we focused on cell-free DNA from human sources in this study. We first investigated the global characteristics of plasma cfDNA in COVID-19 patients. Firstly, cfDNA samples from COVID-19 patients show significantly higher GC content (Fig. 2A) than controls, and the GC contents in COVID-19 patients are positively correlated with IgG levels in the peripheral blood (Supplementary Fig. S1A). Secondly, cfDNA samples from COVID-19 patients show significantly altered size patterns compared to controls. We divided the cfDNA data into short (i.e., < 150 bp), intermediate (150-250 bp), and long (i.e., > 250 bp) categories, as size pattern is a known characteristic that correlates with the tissue origin of cfDNA as well as various physiological conditions of the body (15, 29–31). As a result, cfDNA samples from COVID-19 patients show significantly higher proportions of short fragments (Fig. 2B) while lower proportion of intermediate fragments (Fig. 2C); for the proportions of long fragments, cfDNA from COVID-19 patients do not show significant differences compared to controls; however, mild cases show significantly increased proportion of long molecules than severe patients (Fig. 2D). The cfDNA size pattern is further validated using Bioanalyzer 2100 (Agilent) platform for 10 randomly selected samples (Supplementary Fig. S2). Besides fragment size, end motif pattern is a newly discovered characteristic of plasma cfDNA that correlates with various physiological conditions (32, 33). We analyzed two types of end motifs (termed as 5’-CCCA and CT-5’-CC; see Methods and Supplementary Fig. S1B) in our data. CfDNA samples from COVID-19 patients show significantly increased levels of 5’-CCCA and CT-5’-CC end motif usages than controls (Fig. 2E, Supplementary Fig. S1C). In addition, when 5’-CCCA and CT-5’-CC motif usages are analyzed side-by-side, the COVID-19 blood samples compose two patterns (Fig. 2F, one pattern is highlighted in purple circle). In addition, hypertension is the most common comorbidity in the COVID-19 patients; GC contents and motif usages do not show significant differences between COVID-19 patients with hypertension and without hypertension in the same group, while cfDNA size patterns show slight differences between COVID-19 patients with and without hypertension in the same group (Supplementary Fig. S3). Together, the results demonstrate gross abnormalities in cfDNA characteristics of COVID-19 patients.

**Figure 2.**
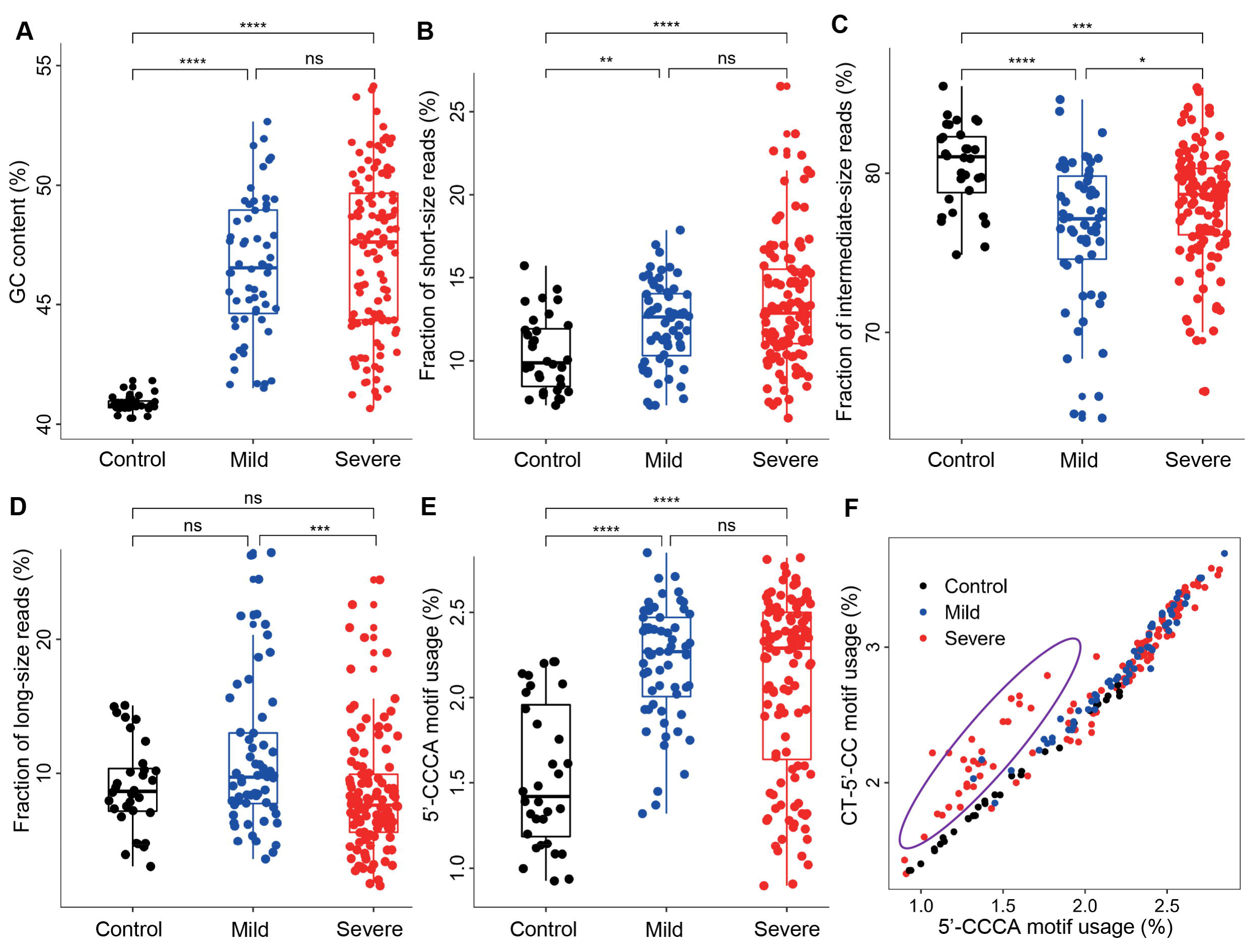
Characteristics of plasma cfDNA in COVID-19 patients. (A) GC content; (B) proportion of short (i.e., < 150 bp), (C) intermediate (i.e., 150-250 bp), and (D) long (i.e., > 250 bp) molecules; (E) proportion of reads with (i.e., usage of) 5’-CCCA end motif; (F) side-by-side comparison of 5’-CCCA and CT-5’-CC end motif usages. In panels A-E, the p-values of statistical comparisons between any groups are shown. ns: non-significant; *: p < 0.05; **: p < 0.01; ***: p < 0.001; ****: p < 0.0001.

### Alterations and dynamics of cfDNA characteristics during treatment

We compared the plasma cfDNA characteristics at the first timepoint (i.e., upon hospitalization) versus the last timepoint, when treatment had taken effect (Fig. 3A-D, Supplementary Fig. S4). COVID-19 patients show significant increase in GC levels after treatment for both mild and severe groups (Fig. 3A). For cfDNA size patterns, differences in proportion of short fragments after treatment are not remarkable in mild patients, while significantly decreased in severe patients; in contrast, both mild and severe groups show significantly elevated proportion of long fragments (Fig. 3B-C). For end motif patterns, elevation in 5’-CCCA and CT-5’-CC end motif usages is marginal in mild patients while significant in severe patients (Fig. 3D). The results thus show that treatment introduces drastic changes to cfDNA characteristics in COVID-19 patients.

**Figure 3.**
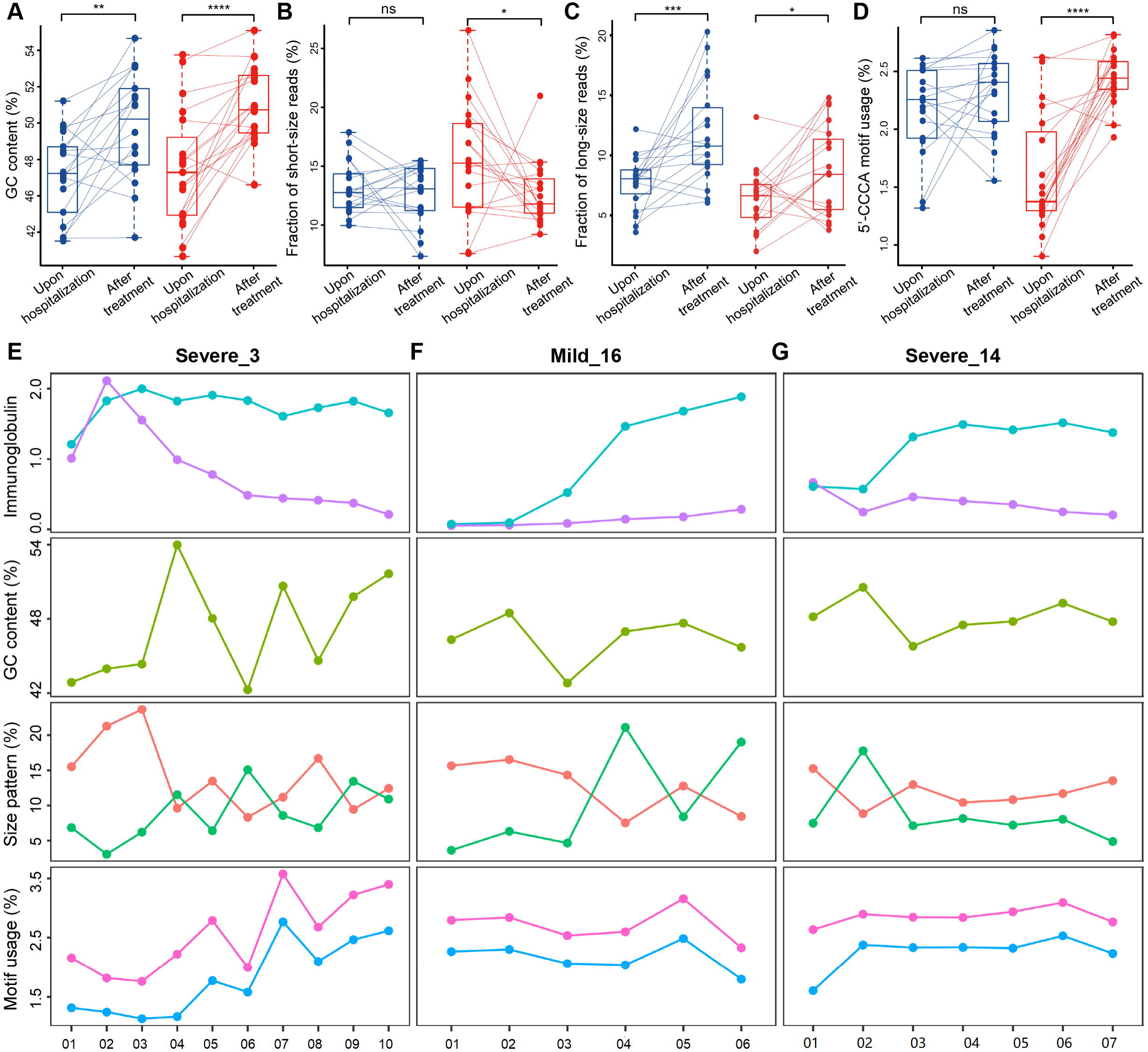
Alterations and dynamics of cfDNA characteristics in COVID-19 patients. (A-D) comparison of GC content, proportion of short/long reads, and usage of 5’-CCCA end motif usage between first (usually upon hospitalization) and last timepoints (when treatment has taken effect) of COVID-19 patients, respectively (dots linked by lines indicate samples from the same patients); (E-G) SARS-CoV-2-specific immunoglobulin levels (Optical Density values), and various cfDNA characteristics during treatment of 3 representative patients. Cyan and purple lines stand for SARS-CoV-2-specific-IgM and SARS-CoV-2-specific-IgG levels, respectively; orange and green lines stand for proportion of short and long fragments, respectively; pink and blue lines stand for CT-5’-CC and 5’-CCCA end motif usages, respectively. The x-axis labels indicate the blood collection date in “Dmmdd” format; for instance, ‘D0127’ means Jan 27^th^, 2020. ns: non-significant; *: p < 0.05; **: p < 0.01; ***: p < 0.001; ****: p < 0.0001.

We further investigated whether cfDNA characteristics could reflect the body responses during treatment. To do this, we profiled cfDNA characteristics along with immunoglobulin levels for COVID-19 patients over the time courses during treatment. Three representative cases (1 mild and 2 severe) are shown in Fig. 3E-G. The SARS-CoV-2-specific IgM level is an important clinical indicator for effective immune response to SARS-CoV-2 infection (3, 34, 35). Hence, for the patient shown in Fig. 3E, the immune system starts to take effect from the second timepoint, when SARS-CoV-2-specific IgG level also starts to increase; however, the other 2 cases (Fig. 3F-G) do not show convincing SARS-CoV-2-specific IgM signal, suggesting possible immune deficiency or insufficient immunization. CfDNA characteristics also show dynamics during treatment in these samples, such as the proportion of long fragments at certain timepoints. In particular, cfDNA end motif patterns gradually increase in the patient shown in Fig. 3E while remain modestly changed in the other two cases.

### Tissue injury signals in cell-free DNA

To explore whether plasma cfDNA could reflect organ damages induced by COVID-19, we adapted our previous orientation-aware cfDNA fragmentation analysis approach (36) to detect signals linked to the tissue origins of cfDNA. Notably, besides blood cells, we focused on lungs, liver, heart, kidneys, pancreas, and brain in this study (Supplementary Tables S1), because these organs are known to be infected by SARS-CoV-2 (6, 7). CfDNA fragmentation patterns for controls are consistent with our previous report that cfDNA coverage decreases in the tissue-specific open chromatin regions if the corresponding tissues contribute DNA in plasma (e.g., blood cells; Fig. 4A), as nucleosome-depletion in such regions makes the DNA unprotected from nuclease digestion (36); however, we find that cfDNA coverage in the open chromatin regions increase in most COVID-19 samples, which may be due to the elevated GC content in cfDNA of COVID-19 patients, as GC content for tissue-specific open chromatin regions are higher than adjacent regions (Supplementary Fig. S5); nevertheless, altered fragmentation signals (e.g., imbalanced coverage patterns) around tissue-specific open chromatin regions are still observed in certain timepoints in almost all severe COVID-19 patients. Fig. 4A shows the coverage signal from the same patients as Fig. 3F-G. For instance, strong fragmentation signals around lung-, pancreas- and brain-specific open chromatin regions are observed at timepoint 2 of the severe case, which echoes the altered cfDNA characteristics (e.g., increase of long fragments) of this patient at the same timepoint (Fig. 3G).

**Figure 4.**
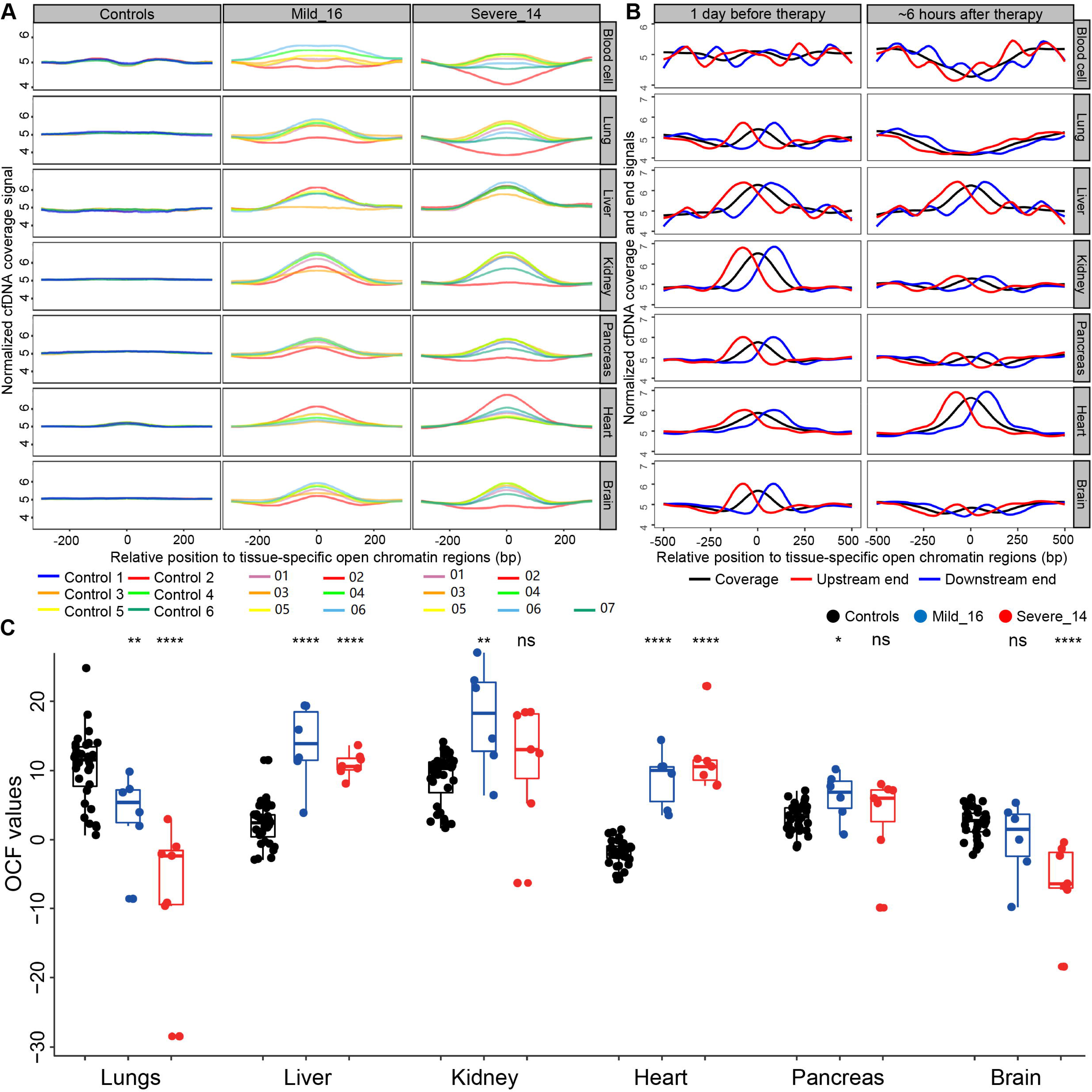
CfDNA fragmentation patterns around tissue-specific open chromatin regions. (A) normalized cfDNA coverage around tissue-specific open chromatin regions in controls, representative mild and severe cases, respectively. CfDNA signals are illustrated in various colors based on the patients or sample collection timepoints. Each row present one tissue and the y-axis show the normalized cfDNA coverage. (B) Plasma cfDNA from 1 day before, and ∼6 hours after treatment of a patient receiving convalescent plasma therapy. Each row present one tissue; y-axis present the normalized read coverage (black line) and orientation-aware end signals (red and blue lines). (C) comparison of OCF values between controls and two representative COVID-19 patients. OCF (<u>O</u>rientation-aware <u>C</u>fDNA <u>F</u>ragmentation) is a measurement approach of cfDNA fragmentation pattern as defined in our previous work (34). Each tissue-of-interest has 3 columns: black, blue, and red dot represents one control, one timepoint in the mild case, and one timepoint in the severe case, respectively. The “ns” and asterisks represent the statistical comparisons between the COVID-19 cases and controls. ns: non-significant; *: p < 0.05; **: p < 0.01; ***: p < 0.001; ****: p < 0.0001.

As an interesting example, we investigated the severe patient who receives convalescent plasma therapy (27, 37) on day 16 of hospitalization. Blood samples are taken 1 day before and ∼6 hours after treatment. Both GC content, size and end motif patterns change remarkably after treatment. Orientation-aware cfDNA fragmentation analysis reveals drastic signal changes after treatment: both coverage and ends around blood cell-, lung-, kidney-, and brain-specific open chromatin regions alter sharply (Fig. 4B). Indeed, clinical records of this patient show various positive changes after treatment that are related to these organs, including returning to normal body temperature and improvements in the lung condition (lesions in the lower right lung field are slightly reduced according to chest radiograph and relief of respiratory distress) as well as consciousness state (increased dose of sedative and muscle relaxant).

Moreover, cfDNA fragmentation patterns for lungs, liver, heart, kidney, pancreas, and brain were quantified using our previous OCF (<u>O</u>rientation-aware <u>C</u>fDNA <u>F</u>ragmentation) approach (see Methods) (36). The results for the two presentative patients illustrated in Fig. 3F-G are shown in Fig. 4C. In general, significantly altered OCF values are observed in the majority of patients and/or tissues, suggesting prevalent tissue injuries in COVID-19 patients. Notably, in COVID-19 patients, OCF values are decreased for lungs and brain, while they are elevated for other tissues. We also observe abnormal OCF values in certain timepoints in the COVID-19 patients while the overall statistical comparisons do not show significant differences (mostly due to limited number of timepoints in this patient or other timepoints show similar OCF values to the controls). To overcome this drawback and to provide explicit tissue injury assessment results, we further built a machine learning-based classification model to predict the tissue injuries based on the orientation-aware cfDNA fragmentation signals (see Methods). The results are summarized in Fig. 5. Notably, clinical diagnoses on tissue injuries for lungs, liver, kidneys, and heart are also available for a proportion of patients. Frequent injuries are observed in various tissues, including lungs, pancreas, and brain, which results are consistent with clinical diagnoses for the majority of patients.

**Figure 5.**
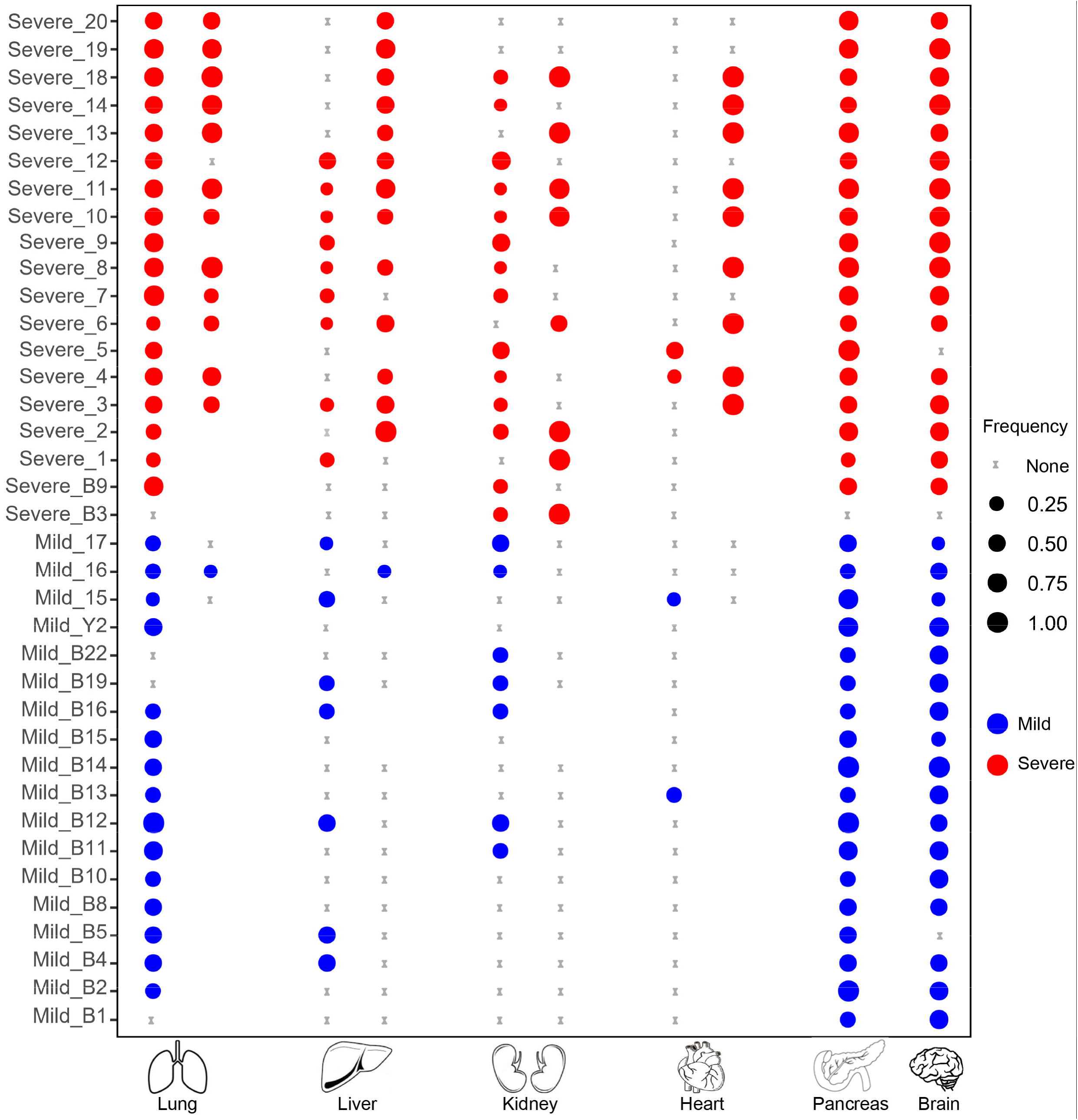
Summary of tissue injury assessment in all COVID-19 patients. For lungs, liver, kidney, and heart, the two columns represent frequencies of cfDNA samples that are predicted to suffer from injuries based on cfDNA fragmentation pattern analysis (left) and clinical diagnoses (right), respectively, for each patient. Blank points mean that clinical diagnoses are not available for these patients. For pancreas and brain, clinical diagnoses are not available for all patients and only the results from cfDNA fragmentation pattern analysis are shown.

## Discussion

The outbreak of COVID-19 has last for more than 1 year. Considering the unclear therapeutics, disease monitoring is of high clinical value for better management and healthcare of the large amount of COVID-19 patients; however, efficient methods are still limited, especially for assessment of various organ injuries (38). In this proof-of-principle study, we have conducted a comprehensive analysis of 176 blood samples collected from 37 COVID-19 patients. Although previous studies on cfDNA characteristics exist, most of them focus on elevated cfDNA concentration and neutrophil extracellular traps (NETs) in the COVID-19 patients (39–42), while our study reveals gross abnormalities and dynamics in a broad range of cfDNA characteristics as well as their clinical potentials in disease monitoring, including increased GC content, altered size and end motif patterns (Fig. 2A-D). COVID-19 patients suffer from active immune response to the viral infection and produces high level of immunoglobulins (35, 43), which prefer binding/protecting GC-rich DNA (e.g., DNA molecules originated from the open chromatin regions) (44), suggesting that immune response may be responsible to the abnormalities in plasma cfDNA characteristics in COVID-19 patients. Moreover, the NET process is known to generate long cfDNA molecules; Fig. 2D shows that mild COVID-19 patients tend to have more long cfDNA molecules than controls while severe patients do not. This result suggests that patients in the mild group may have a higher innate immune activity than those in the severe group, which may explain why their symptoms are weaker. In the meantime, Fig. 3C shows that after treatment, the proportions of long cfDNA molecules are increased in both mild and severe COVID-19 patients, suggesting more NETs, i.e., enhanced immune responses of the patients after treatment, which is consistent with the improvement of clinical symptoms in these patients. It is also interesting to see differences in size patterns between COVID-19 patients with and without hypertension (Supplementary Fig. S3), as previous studies have demonstrated that cfDNA alterations could serve as a diagnostic biomarker for cardiovascular diseases (45). In addition, end motif analysis reveals two patterns in COVID-19 patients; interestingly, most of the samples that form the altered pattern (Fig. 2F, purple circle) are collected at the first or second timepoints of severe patients, when the patients’ conditions are most critical (e.g., in a coma). Plasma cfDNA fragmentation patterns could be affected by various biological and clinical scenarios, while current knowledge in this field is still limited. Hence, the altered cfDNA signals may suggest aberrant, yet elusive, cell death in COVID-19 patients.

Furthermore, plasma cfDNA reveal disease dynamics and organ injury signals during the treatment. For instance, significant changes are observed in cfDNA samples at the last timepoint compared to the first timepoint (Fig. 3A-D), indicating that cfDNA characteristics could reflect therapeutic efficacies. Moreover, cfDNA show fragmentation signals around tissue-specific open chromatin regions in various cases, which is partly in line with clinical observations on organ injuries in these patients. In fact, organ injury in COVID-19 patients may correlate and partially explain the altered characteristics in cfDNA, as cells in damaged organs may die abnormally thus release DNA with aberrant fragmentation patterns (Fig. 2) (46). As an interesting example, cfDNA from a severe case receiving plasma therapy show huge alterations ∼6 hours after treatment: we observe drastic changes around blood cell- and lung-specific open chromatin regions, suggesting that the patient has responded to the treatment, especially the lungs, which is evidenced by the clinical observations; kidney-specific open chromatin regions also show strong fragmentation patterns after treatment, which is reasonable because kidney is an important organ for metabolism and is known to involve in COVID-19 (47). Hence, the data indicate that cfDNA analysis is indeed sensitive in monitoring the body response during treatment.

Detection and monitoring of organ injuries are highly valuable for COVID-19 patients. Tissue injury assessment could be indicative for potential sequelae of the patients as COVID-19 patients frequently suffer from multiple tissue injuries even months after discharge (11), and organ failure is a major cause of mortality in COVID-19 (25, 48). In this study, we compared the quantified orientation-aware fragmentation patterns (i.e., OCF values) between COVID-19 patients and controls (Fig. 4C), which results show frequently altered fragmentation patterns in COVID-19 patients, suggesting that tissue injuries are indeed common in COVID-19 patients. Interestingly, the OCF values for lungs and brain show an opposite direction in COVID-19 patients compared to other tissues. The underline mechanisms are elusive; while for lungs, we think that it may be related to their unique position as the primary organ of viral infection where frequent non-apoptotic cell deaths may occur. Besides the statistical comparisons, we further developed a machine learning-based approach to for qualitative (i.e., yes or no) measurement of tissue injuries, which could provide an explicit result for easier interpretation of the data.

Considering that cfDNA analysis could reveal tumor signals long before clinical diagnosis (49), we think that cell-free nucleic acid analysis could be more sensitive than clinical diagnosis in tissue injury assessment. For instance, cfDNA analysis shows that almost all COVID-19 patients suffer from lung injury which is consistent with the fact that lungs are the primary infection sites in COVID-19. Besides lungs, kidneys, pancreas, and brain are other organ with frequent injuries, which is consistent with clinical reports on COVID-19 (50–52). Hence, COVID-19 induced low-level oxygen in the blood, blood clots, and cytokine storms can cause kidneys to malfunction (53); diabetes is one of the most common comorbidities in COVID-19 patients and COVID-19 also causes diabetic symptoms in the non-diabetic patients (54, 55); neurological abnormalities are also common in COVID-19 patients (56, 57). Furthermore, currently convalescent plasma therapy is a legal, yet controversial, therapeutic method for COVID-19 (27, 58). Through analyzing the blood sample from one patient with plasma therapy, we show that although plasma therapy makes improvements of clinical symptoms in this patient, it also introduces various tissue injuries therefore suggesting that dedicated medical inspections on various organs would be helpful for patients receiving plasma therapy.

In the meantime, we only have limited clinical data for tissue injury assessment in the COVID-19 patients, and clinical diagnoses for pancreas and brain are not available at all. In fact, clinical approaches for organ injury assessment usually require dedicated assays for assessment of each tissue, while such assays may not be feasible, or with a low priority, when the medical system is overloaded during the outbreak of the pandemic; as a contrast, cfDNA is much more favorable as it able to profile the injury landscape of various organs from one tube of peripheral blood, therefore promises a much more efficient and convenient approach. In previous studies, Cheng et al. and Andargie et al. have also investigated tissue injuries in cfDNA through tissue-specific DNA methylation markers (59, 60); however, considering the experimental challenges and complexities in current cfDNA methylation profiling assays, our approach significantly lowered the experimental difficulty as we only require routine whole-genome sequencing. In particular, the dynamics of cfDNA characters and tissue injury signal for a mild and a severe patient (Fig. 3, 4) show favorable consistency (e.g., kidney injury signal in the mild case, and signals of multiple tissue injuries in the severe case), demonstrating the potential of cfDNA in disease monitoring during treatment.

On the other hand, there are various limitations in this study. Firstly, the clinical diagnosis for many COVID-19 patients and tissues are not available due to the limited medical resources during the outbreak of the pandemic. Secondly, we could only perform qualitative analyses without comprehensive statistical analyses for tissue injuries. Hence, to provide more meritorious information to the clinic, it is worthwhile to validate the results using larger and more thorough datasets in the following studies. In addition, it would be favorable to explore the feasibility of other analyses, such as nucleosome positioning (14, 36) and promoter coverage patterns (61) , for quantitative measurement of organ injuries in following works.

As a summary, through analysis of cfDNA in COVID-19 patients, we report alterations and dynamics of cfDNA characteristics during treatment, as well as organ-specific signals in cfDNA, demonstrating that cell-free DNA could serve as valuable analytes for effective disease monitoring and tissue injury assessment of COVID-19 patients.

## Methods

### Ethics approval and patient recruitment

This study had been approved by The First Affiliate Hospital of Guangzhou Medical University Ethics Committee, and the institutional review board of BGI; written informed consents had been obtained from all patients and healthy donor participated in this study. A total of 37 COVID-19 patients and 32 non-COVID-19 controls were recruited from local hospitals in Guangdong. The COVID-19 patients were divided into mild (N=18) or severe (N=19) groups according to the Guidelines for COVID-19 Diagnosis and Treatment (Trial Version 5) (27) issued by the National Health Commission of China. Control subjects were collected from the same hospitals as the COVID-19 patients based on the following criteria: negative for SARS-CoV-2 tests on the blood-taken day and has never been diagnosed to have COVID-19 until the end of this study, and comparable age distribution to the COVID-19 patients. Blood samples were collected during Jan 27 to Mar 28, 2020.

### Clinical data acquisition and analysis

The epidemiological, demographic, clinical, laboratory characteristics and treatment data were extracted from electronic medical records, and all the data had been double-checked by the relevant physicians to ensure the accuracy and completeness of the epidemiological and clinical findings. Frequency of clinical examinations was determined by the physicians-in-charge.

Diagnoses of severe pneumonia and ARDS (Acute Respiratory Distress Syndrome) in the COVID-19 patients were according to Diagnosis and Treatment Protocol for Novel Coronavirus Pneumonia (Trial Version 5) (27) and the Berlin Definition (62), respectively. Kidney injury was diagnosed according to the Kidney Disease: Improving Global Outcomes (KDIGO) guideline (63). Heart injury was diagnosed if serum levels of cardiac biomarkers (e.g., cardiac troponin I) were above the 99th percentile upper reference limit, or if new abnormalities were shown in electrocardiography and echocardiography (25). Liver function indicators measured on admission, including alanine aminotransferase (ALT), aspartate aminotransferase (AST), direct bilirubin, etc.; patients whose ALT or AST is above the normal range were considered to suffer from liver function abnormality (64). Pancreatic function tests were not carried out for most patients in our cohort; in addition, most patients are in a state of sedation and neurologic examinations (e.g., brain MRI) were also omitted (57).

### cfDNA extraction and processing

All blood samples (including those from the controls and COVID-19 patients) are collected and processed according to consensus guideline for cell-free DNA analysis (65). Briefly, for each sample, 1ml peripheral blood was collected using EDTA anticoagulant-coated tubes, then centrifuged at 1,600g for 10 min at 4°C within six hours after collection; the plasma portion was harvested and recentrifuged at 16,000g for 10 min at 4 °C and to remove blood cells. Cell-free DNA was extracted from 200 µl plasma using MagPure Circulating DNA KF Kit (MD5432-02, Magen) following the manufacturers’ protocols. Sequencing libraries was prepared using MGIEasy Cell-free DNA Library Prep kit (MGI) on the amplified cfDNA following the manufacturer’s protocol. All the cfDNA libraries passed quality control and sequenced on DNBSEQ platform (BGI) in paired-end 100 bp mode.

### CfDNA sequencing and data processing

We used SOAPnuke (v1.5.0) (66) software to trim sequencing adapters, filter low quality and high ratio Ns in the raw reads with default parameters. The preprocessed reads were then aligned to the human reference genome (NCBI build GRCh38) using BWA (67) software with default parameters. After alignment, PCR duplicates were removed using in-house programs: if more than two reads shared the same start and end positions, only one was kept for following analyses and the others were discarded as PCR duplicates.

### CfDNA characteristics profiling

For each cfDNA sample, GC content was determined as the proportion of G or C in the sequenced nucleotides; fragment size for each molecule was determined as the distance between the two outmost ends obtained from the alignment result; short fragments were defined as reads shorter than 150 bp, and long fragments were defined as reads longer than 250 bp. Considering that most nucleases in mammals function in an endonuclease manner (i.e., they bind to DNA and cut within the bound sequence), besides the 4-mer motifs at the 5’-end of cfDNA as in previous studies (32, 33), we extended 2 bp from the 5’-end and proposed a novel 4-mer motif definition: 5’-CCCA motif usage was calculated as the proportion of reads starting with CCCA, and CT-5’-CC motif usage was calculated as the proportion of reads starting with CC and the 2 bp in the genome prior to the 5’-end are CT. The definition of 5’-CCCA and CT-5’-CC motifs are illustrated in Supplementary Fig. S1B. As a result, the previous definition presents CCCA while our new definition reveals CTCC as the motif with the highest usage. Notably, in our cohort, the CT-5’-CC motif usage is positively correlated with, and always higher than, 5’-CCCA, suggesting that our newly discovered CT-5’-CC motif could also reflect enzymatic preferences during cell apoptosis.

### Orientation-aware cfDNA fragmentation analysis

In our previous work (36), we had mined and investigated tissue-specific open chromatin regions for blood cells, lungs, liver, intestines, breast, ovary, and placenta. Based on clinical reports on tissue injuries of COVID-19 patients (6), we added kidney, pancreas, heart, and brain into the tissue list, while removed placenta from the tissue list (as there is no pregnancy samples in our cohort) in the current study. Tissue-specific open chromatin regions for all the tissues in the list were mined using the same algorithm as described in our previous work. The accession numbers of the Dnase I hypersensitivity data and the final list of tissue-specific open chromatin regions used in this study were summarized in Supplementary Table S1. For each cfDNA sample, coverage and end pattern around the tissue-specific open chromatin regions were profiled using the same algorithm as described in our previous work (36). To minimize the biases of the abnormally high coverage in the center of open chromatin regions in COVID-19 patients (Fig. 4A), OCF values for each patient and tissue were quantified using (−210, −180) and (180, 210) windows around the tissue-specific open chromatin regions.

### Prediction of tissue injury using cfDNA fragmentation pattern

Considering that the GC content is significantly elevated in COVID-19 samples (Fig. 2A), to minimize the potential biases (e.g., from sequencing), we developed a new method to infer tissue injury signals that solely depends on the cfDNA data from the COVID-19 samples. Based on the knowledge that blood cells are the major contributor of cfDNA in most clinical scenarios (19, 68) and to date there is no clinical/genetic evidence of ovary injuries in COVID-19 patients (in fact, a large proportion of the COVID-19 patients are male in our cohort), we utilized the orientation-aware cfDNA fragmentation pattern around blood cell- and ovary-specific open chromatin regions from all COVID-19 blood samples as positive and negative signals, respectively, to train a classification model for injury assessment of other tissues. Briefly, for each cfDNA sample, after profiling of orientation-aware cfDNA end signals around the tissue-specific open chromatin regions, for all the tissues-of-interest (i.e., blood cell, ovary, lungs, liver, kidneys, pancreas, heart, and brain), the differences in normalized upstream (*U)* and downstream (*D)* end signals were calculated for each locus in two symmetrical 30 bp windows around the corresponding tissue-specific open chromatin regions (i.e., (−210,−180) and (180,210)); hence, a vector of 60 values would be obtained for each tissue; then, we collected all the vectors for blood cells and ovary in the COVID-19 blood samples as positive and negative datasets, respectively, to train a classification model using SVM (Support Vector Machine) approach (69). During training, a 5-fold cross-validation was employed, which showed an overall accuracy of 93.5% on the training dataset. After model-training, for each of the tissue-of-interest, we applied the SVM classification model on its *U* and *D* end signal difference vector to determine whether it showed injury or not, during which procedure a score (calculated by the classification model) of 0.8 was used as the classification cutoff. Lastly, for each patient, we calculated the frequency of positive injury predictions in his/her blood samples for all the tissues-of-interest as the final prediction results (Fig. 5).

### Statistical analysis

Comparisons of cfDNA characteristics between COVID-19 patients and controls were performed using Mann-Whitney *U* test; comparisons of cfDNA characteristics for COVID-19 patients at the first and last timepoint were conducted using Wilcoxon signed-rank test; comparisons between OCF values for COVID-19 patients and controls were performed using Mann-Whitney *U* test. All p-values are two-tailed and a p-value lower than 0.05 was considered as statistically significant.

### Data access

The data that support the findings of this study have been deposited into CNGB Sequence Archive (CNSA)(70) of China National GeneBank DataBase (CNGBdb) (71) with accession number CNP0001306.

## Data Availability

Data access
The data that support the findings of this study have been deposited into CNGB Sequence Archive (CNSA)(70) of China National GeneBank DataBase (CNGBdb) (71) with accession number CNP0001306.

https://db.cngb.org/cnsa/project/CNP0001306/reviewlink/

## Author contributions

X Jin, J Zhao, K Sun, F Cheng, and Y Xu designed the study; Y Wang, F Cheng, R Ou, Y Li, H Zhou, S Lin, F Xiao, Y Jin, F Zheng, L Zhang, Z Zhang, A Zhu, J Zhao, Y Li, L Wen, F Li, Y Zhang, J Sun, Z Zhuang, Z Chen, J Zhuo and S Zhang collected clinical specimen, summarized clinical data and performed experiments; J Xu, K Sun, X Jin, Y Lin, Y Luo, Z Zhang, W Chen, A Zhu, T Kuo, J Zhao, L Lin, Y Tao, Y Li, L Li, L Wen, H Yuan, H Sun, J Li, and F Li analyzed and/or interpreted data; W Chen, J Xu, Y Luo, Y Lin, Y Wang, F Cheng, Y Jin, F Zheng, Y Li, H Zhao, S Lin, F Xiao and L Zhang analyzed clinical data; K Sun, J Xu, Y Wang, Y Luo, and W Chen drafted the manuscript; X Jin, J Zhao, K Sun, Y Xu, N Zhong, X Xu, J Wang, H Yang, P Wei, J Yuan, T Xu, D Chen, and Y Sun revised the manuscript critically for important intellectual content; X Jin, K Sun and J Zhao supervised the project. All authors had read and approved the manuscript. All authors agreed to submit the manuscript, read and approved the final draft and take full responsibility of its content, including the accuracy of the data and its statistical analysis.

## Acknowledgments

We would like to thank Ms. Qi Wang from Shenzhen Bay Laboratory for her technical assistance. This study has been supported by China National GeneBank (CNGB), Shenzhen Bay Laboratory, National Key Research and Development Program of China (2018YFC1200100 to JZ), National Science and Technology Major Project (2018ZX10301403 to JZ), the emergency grants for prevention and control of SARS-CoV-2 of Ministries of Science and Technology, and Education of Guangdong province (2020A111128008, 2020B1111320003, 2020KZDZX1158 to JZ, and 2020B1111330001 to NZ), National Natural Science Foundation of China (32000398 to XJ), Natural Science Foundation of Guangdong Province, China (2017A030306026 to XJ), Guangdong-Hong Kong Joint Laboratory on Immunological and Genetic Kidney Diseases (2019B121205005 to XJ), Guangdong Provincial Key Laboratory of Genome Read and Write (No. 2017B030301011 to XX), Guangdong Provincial Academician Workstation of BGI Synthetic Genomics (No. 2017B090904014 to HY), and BGI-research (BGIRSZ2020007 to KS).

