## Supplementary figures and images for "Plasma cell-free DNA promise disease monitoring and tissue injury assessment of COVID-19"

### Supplementary Fig S1

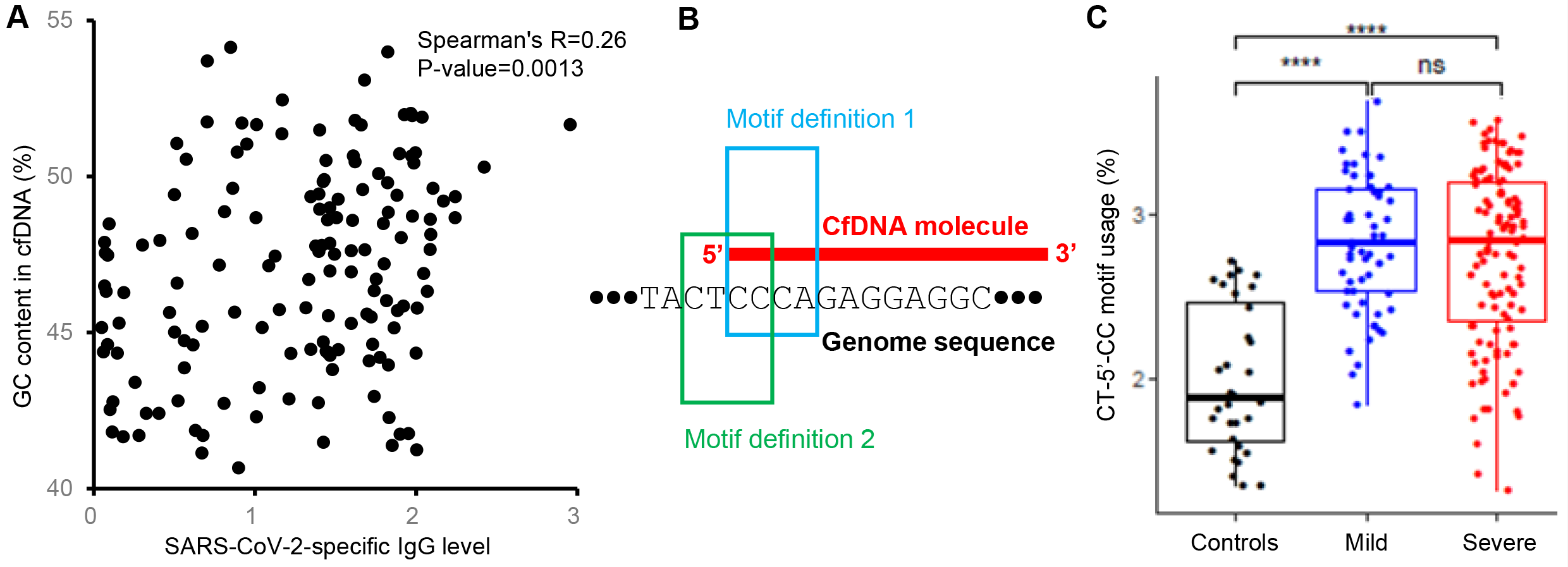

### Supplementary Fig S2

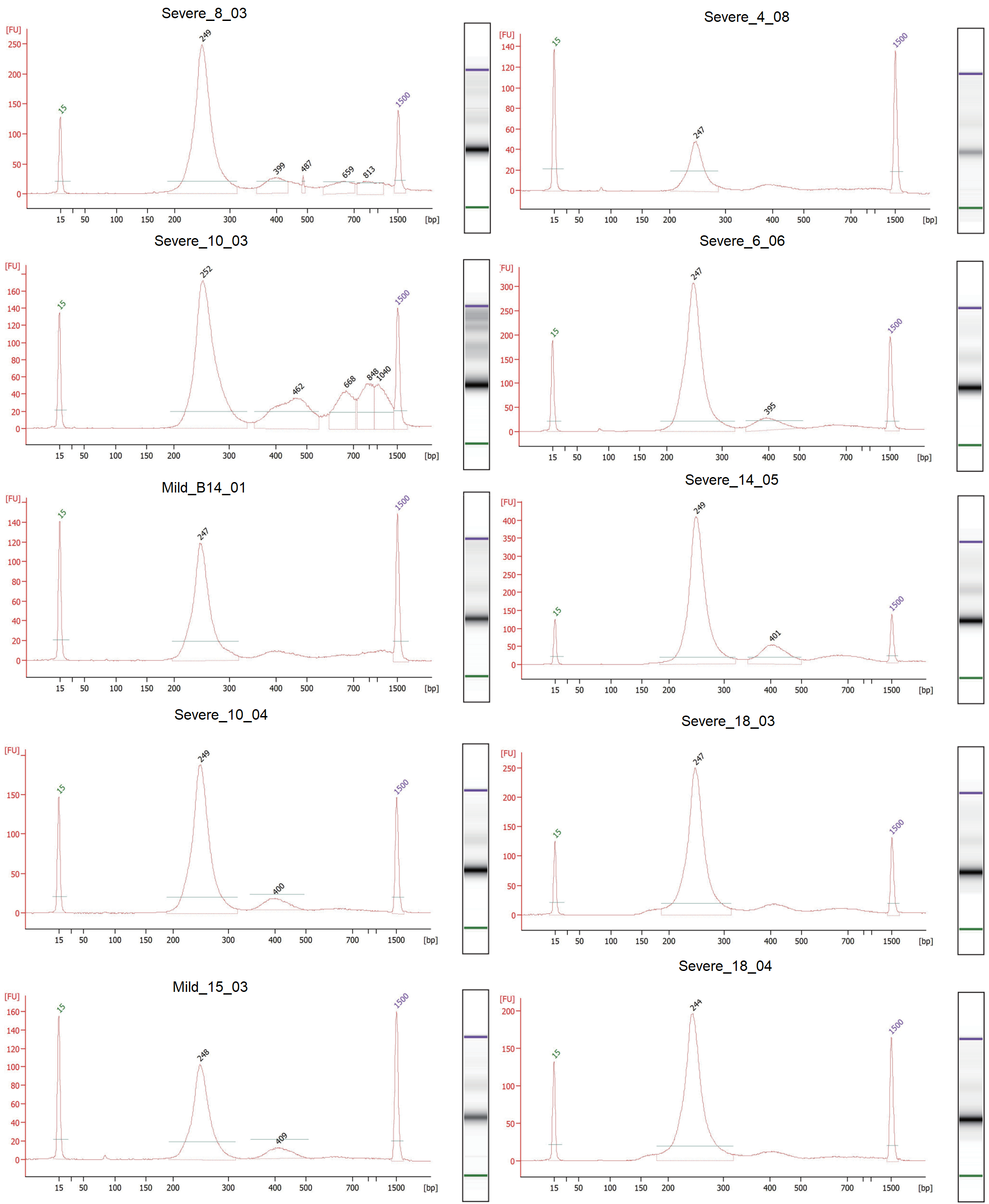

### Supplementary Fig S3

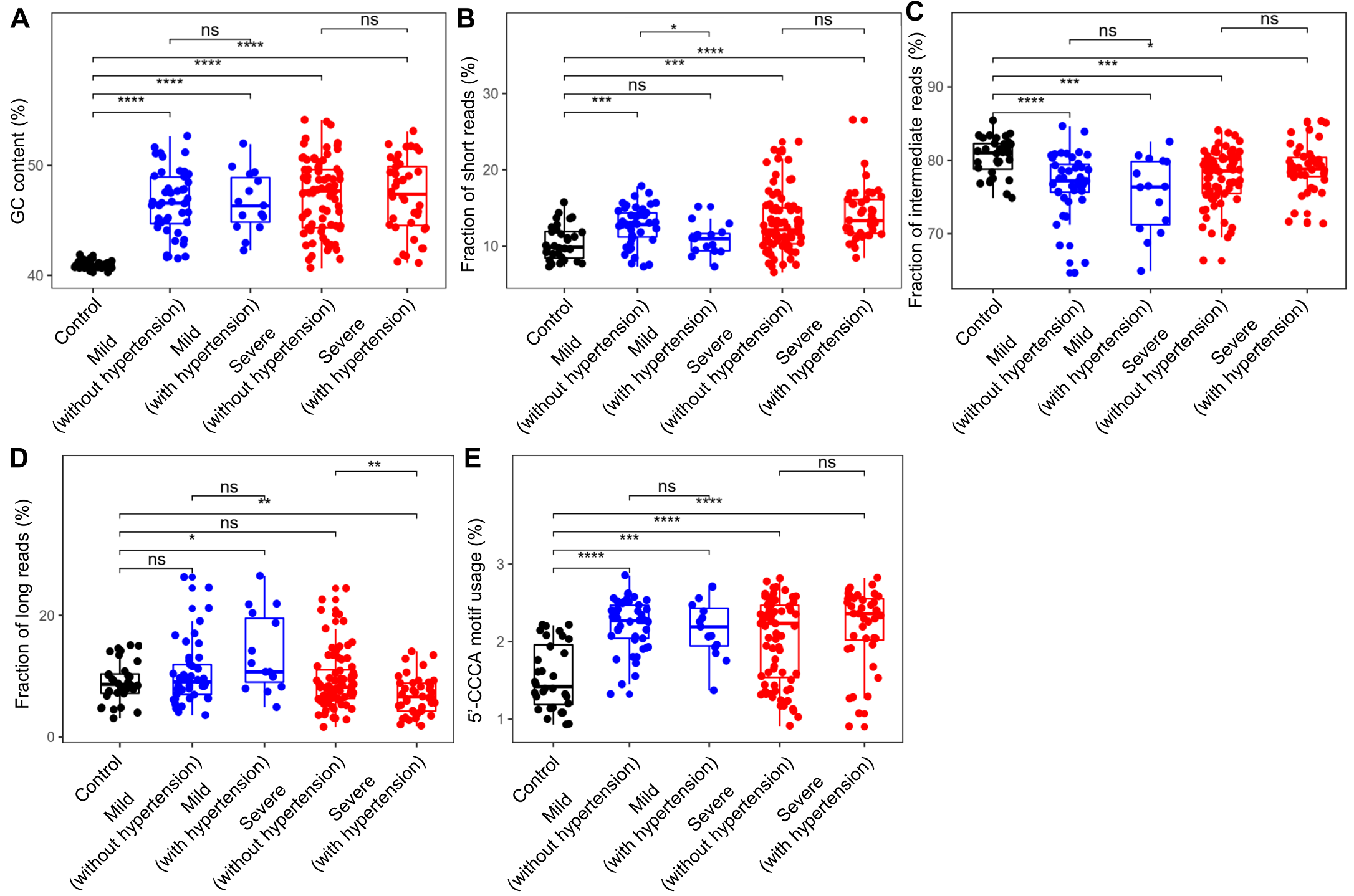

### Supplementary Fig S4

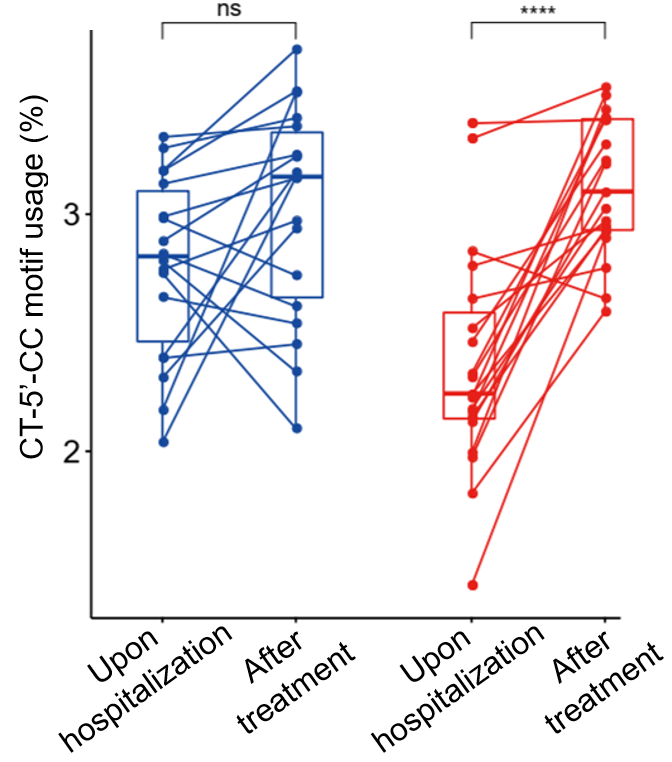

### Supplementary Fig S5

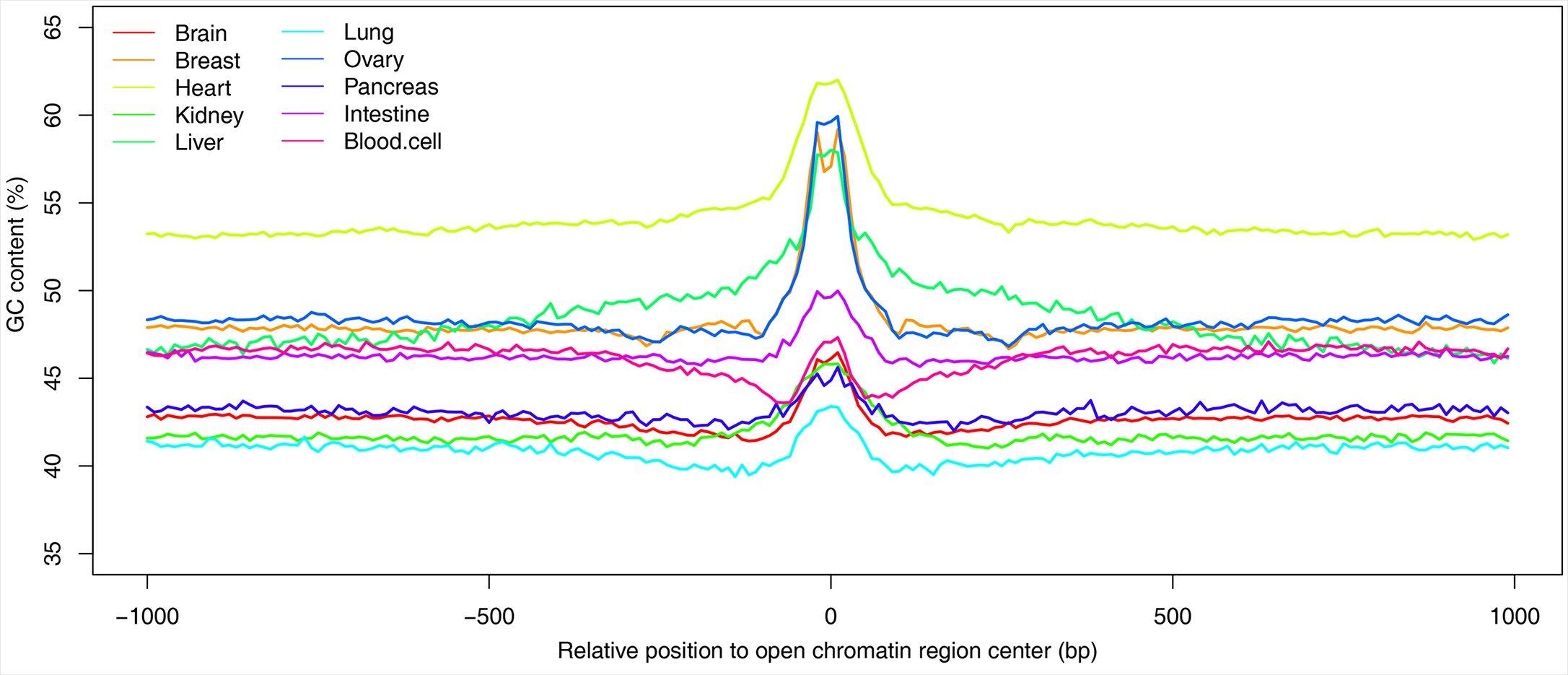
